# Clinical epidemiology of intrapartum stillbirths in The Gambia and Burkina Faso: Descriptive cohort analysis from PregnAnZI-2 clinical trial

**DOI:** 10.1101/2025.10.22.25337328

**Authors:** Fatoumata Sillah, Toussaint Rouamba, Helen Brotherton, Christian Bottomley, Bully Camara, Joël D Bognini, Usman N Nakakana, Guétawendé JW Nassa, Nathalie Beloum, Edmond Yabré Sawadogo, Athanase M. Somé, Chiquita Jones, Shashu Graves, Madikoi Danso, Yusupha Njie, Ebrahim Ndure, Umberto d’Alessandro, Halidou Tinto, Anna Roca

## Abstract

**Introduction:** West Africa has the highest intrapartum stillbirth rate globally, yet there is limited understanding of regional clinical and epidemiological risk factors. Local data on intrapartum stillbirth rates and risk factors is urgently needed to address the knowledge gap, identify vulnerable sub-groups, and inform policy to reduce stillbirth rates in West Africa.

**Methods:** This descriptive cohort study is a secondary analysis of data collected in the framework of a randomised clinical trial, conducted between 2017 and 2021 in The Gambia and Burkina Faso. Pregnant women considered to be low-risk for intrapartum complications and their offspring were enrolled during labour at ten primary and secondary health facilities in The Gambia and Burkina Faso. Intrapartum stillbirth was detected prospectively. Associations between independent clinical and socio-demographic factors and intrapartum stillbirth were determined using logistic regression models, informed by a novel conceptual framework.

**Results:** 11,983 women and their 12,198 offspring were included in this analysis. We identified 93 intrapartum stillbirths, representing a rate of 7.6/1000 total births. Pre-conception and antepartum risk factors for intrapartum stillbirth were identified as: parity (*p*=0.02), with highest risk for primiparous women compared to parity of 2 (aOR 4.16, 95% CI 1.71-10.14); history of previous stillbirth (aOR 8.75, 95% CI 4.88 - 15.67) or previous caesarean section (aOR 4.52, 95% CI 1.53 - 13.33); maternal antibiotics before labour (aOR 4.67, 95% CI 1.10-19.86); twin births (aOR 3.53, 95% CI 1.73 - 7.21); and extremes of birth weight, for both macrosomia (aOR 4.97, 95% CI 1.74-14.17) and low birth weight (LBW)(aOR 1.91, 95% CI 1.07-3.40).

***C*onclusion:** This study confirms that intrapartum stillbirths are a major public health problem in rural and urban West Africa. Early identification of at-risk women and pregnancies (e.g., history of previous stillbirth or caesarean section, twins, foetal macrosomia, or LBW) with enhanced antenatal monitoring and planning for safe delivery could reduce intrapartum stillbirth rates in the region.

## INTRODUCTION

An estimated 2 million stillbirths (newborn showing no signs of life at birth) occur globally every year.(1, 2) The global stillbirth rate (SBR) in 2021 was 13.9 per 1000 total births,(1) a rate comparable to the overall neonatal mortality rate.(3) As with other adverse perinatal outcomes, the burden of stillbirths disproportionally affects low- and middle-income countries (LMIC), particularly sub-Saharan Africa (SSA) and southern Asia where three-quarters of all stillbirths occur.(1) The contribution of SSA to the global burden of stillbirths has increased from 27% to 42% over the last two decades, with an overall SBR of 21.7 per 1000 total births in 2019, compared to 3.1 per 1000 total births in Europe and North America.(2) If this trend persists, SSA may account for half of all global stillbirths by 2030.(2) Despite the substantial impact of stillbirths on mothers, families, communities, and health systems,(4, 5) research on this area has been historically neglected.(2, 6) However, recent global public health efforts to prevent stillbirths have recently increased due to the Every Newborn Action Plan, a road map of evidence-based perinatal strategies aiming for all countries to reduce the national SBR to <12 per 1000 total births by 2030.(7, 8)

Around half of all stillbirths in SSA occur intrapartum,(2) defined as after the onset of labour and prior to delivery,(1, 9) compared to 6% in Europe and North America.(2) There is growing recognition that antenatal (before labour) and intrapartum stillbirths involve different causal pathways,(10) hence it is important to consider them separately. West Africa, one of the poorest regions in the world, has countries with the highest intrapartum stillbirth rates, for example Cote d’Ivoire (14 per 1000 births) and Nigeria (11 per 1000 births), compared to intrapartum SBR of 1.2 per 1000 births in HIC settings.(11) Over two-thirds (68%) of deliveries in West Africa occur in health facilities,(12) offering opportunities to detect complications and provide targeted interventions to prevent intrapartum stillbirth in this region. Despite a growing body of literature focused on understanding risk factors for stillbirths in SSA, previous studies frequently lacked adjustment for confounding factors and did not distinguish between antenatal and intrapartum stillbirths.(13) A recent systematic review reported determinants of >20,000 stillbirths from 37 studies in SSA, yet only four West African countries (Ghana, Liberia, Sierra Leone and Nigeria) were represented, with two population based West African studies, both in Ghana.(13) This systematic review also highlighted the current lack of evidence to inform understanding of the interplay between individual-level risk factors and health system risk factors in varying SSA contexts, important to sustainably address underlying root causes of stillbirths.

The epidemiology and risk factors for stillbirths differ between South-East Asia and SSA(10) and likely also vary within and between heterogeneous SSA sub-regions. A comprehensive understanding of risk factors for intrapartum stillbirths in varying regions would enable development of context specific prediction models suitable for use within existing resource limited health systems.(14) The study presented here aims to describe the incidence and risk factors for intrapartum stillbirths, including consideration of factors related to health-facility delivery, in The Gambia and Burkina Faso, two West African countries with current data gaps for this important public health issue.

## METHODS

### Study design

This study comprises of secondary data analyses from the PregnAnZI-2 trial, a phase III, double-blind, placebo-controlled individually randomised clinical trial in which ∼12,000 women in The Gambia and Burkina Faso received either intra-partum azithromycin or placebo.(15) The PregnAnZI-2 trial did not identify evidence of the intervention’s effect on stillbirths,(16) hence the whole cohort are included in this post-hoc analysis.

### Study setting

Data was collected between October 2017 and May 2021 from women who delivered at one primary and one secondary health facility in The Gambia and eight primary health facilities in Burkina Faso. Sites were selected to represent both urban and rural health systems and populations, with findings generalisable to other West African countries. Estimates from mathematical models suggest that the population-level total SBR in 2021 was approximately 21 per 1,000 births in The Gambia and 20 per 1,000 births in Burkina Faso.(1)

The two Gambian trial sites were in the urban western region: Serekunda Health Centre (SHC) and Bundung Maternal and Child Health Hospital (BMCHH). During the trial period the sites collectively conducted over 7,000 deliveries/year. SHC provided basic antenatal and intrapartum care. All women requiring emergency obstetric care, including caesarean section, were referred to Kanifing General Hospital, a secondary level facility located 3.4 kilometres (km) away. BMCHH provided emergency obstetric care and only rarely were women referred to the tertiary level Edward Francis Small Teaching Hospital in Banjul, located approximately 15 km away.(17)

All eight sites in Burkina Faso were primary health facilities located in the rural districts of Nanoro and Yako.(16) Emergency obstetric care was not available locally and women requiring emergency caesarean section were referred to the Centre médical avec antenne chirurgicale (CMA) de Nanoro or CMA de Boussé, located maximum 25 km away. If a woman delivered somewhere other than the trial catchment area, this was recorded as “other” and this was predominantly at a referral hospital where specialised expertise and emergency obstetric care were available.

The climate in both countries is typical of the sub-Sahel region, with a long dry season from November to May and a short rainy season between June and October. The population of the catchment areas are representative of both countries, including their main ethnic groups, Mandinka (The Gambia) and Mossi (Burkina Faso).

### Participant eligibility and recruitment

This study included all pregnant women recruited to the PregnAnZI-2 trial and their offspring. Inclusion criteria for study women were age >16 years and delivery at a study site. Women were excluded from PregnAnZI-2 trial if any of the following criteria were present: in-utero foetal demise (detected as absence of foetal heart beat) when attending the study health facility during labour; planned caesarean section delivery; antenatal diagnosis of severe congenital malformation; known acute and/or chronic condition before enrolment, including HIV infection; known macrolide allergy or maternal use of drug known to prolong QT interval during preceding two weeks (e.g. erythromycin, chloroquine).(15) Most enrolled pregnant women were apparently healthy at enrolment. However, despite clear exclusion criteria, it was not possible to exclude all women with an acute or chronic medical condition due to health system limitations at the trial sites and lack of rigorous diagnosis of antenatal and intrapartum complications.

### Study procedures

Written informed consent was obtained antenatally and verbally confirmed when the woman presented at the trial health facility in labour.(15) Eligibility was checked during enrolment, including confirmation of labour and measurement of foetal heart rate by the study and/or health facility midwife using a hand-held foetal doppler, provided and maintained by the study team.(15)

All obstetric and immediate newborn care, including resuscitation, was provided and led by health facility staff with the presence of a research nurse at delivery who assisted when needed and/or a research clinician if available. Newborns who were unable to initiate and sustain adequate respirations immediately after delivery were resuscitated by drying and providing warmth, oral suctioning, bag-valve-mask ventilation, chest compressions and oxygen therapy via nasal prongs, according to the Helping Babies Breathe protocol of resuscitation.(15) Occurrence of stillbirth was identified on clinical assessment by the attending facility clinical staff and documented immediately thereafter by both the research nurse present at the delivery and the research clinician on duty. Neonates, including stillbirths, were weighed at birth or within 24 hours, using digitally calibrated electronic weighing scales (SECA 384/385) to 0.1kg precision. Socio-demographic, antenatal and intra-partum clinical data were collected by a research nurse prior to maternal hospital discharge using direct observation, questioning of mother and source documents (antenatal cards, medical records and partographs).(15)

### Definitions and outcome measures

There is no universally accepted definition of stillbirth, with variations in gestational age, weight and crown-heel length thresholds depending on global region and reporting purpose.(9) In this study we used the ICD-11 definition of stillbirth: complete expulsion, or extraction, of a foetus following its in-utero demise at gestational age >22 weeks.(18) This was chosen to avoid restrictive gestational age cut-offs in view of absent accurate gestational age estimates for the majority of pregnancies in our cohort. Intrapartum stillbirths are typically described as being fresh, referring to intact skin with no disintegration, which are assumed to occur <12 hours before delivery.(19) Macerated stillbirths (skin oedema or breakdown) are typically associated with antenatal death due to prolonged presence in-utero, but can also occur during the intrapartum period if labour is very prolonged. As enrolled women were in established labour and had a foetal heart rate checked prior to recruitment, all stillbirths in this cohort are by definition intrapartum (foetal demise after onset of labour and prior to delivery),(9) including both fresh and macerated stillbirths. We did not document duration of labour. The outcome of each delivery was categorised as either a livebirth or stillbirth by health facility personnel according to absence of signs of life (e.g., absence of breathing and heartbeat) with presence of skin or soft tissue changes used to identify macerated stillbirths.

### Data management and statistical analysis

Data was entered into an electronic database on the REDCap platform using encrypted and reliable mobile devices and computers at the study sites. All data were pseudo-anonymised with unique identification numbers. Consistency checks and data validation were performed at regular intervals.(15)

Statistical analyses were conducted using Stata version 17.0 (Statacorp). The intrapartum SBR was calculated as the number of stillbirths per 1000 total births.(1) As intrapartum stillbirth is the definitive adverse outcome for intrapartum related asphyxia (IRA) and these two conditions exist on a spectrum, a conceptual framework to understand pathways to IRA(20) was adapted to visualise potential variables associated with intrapartum stillbirth, including consideration of health system factors (figure 1).(13) This framework was used to select independent variables for unadjusted and adjusted logistic regression to identify risk factors for intrapartum stillbirth versus live birth, expressed as crude and adjusted odds ratios (OR) respectively, with 95% confidence intervals (CI) to represent degree of certainty, and *p* values. For categorical variables, the baseline category was chosen to reflect either a “normal pregnancy and delivery” (e.g., normal birth weight, non-extremes of parity) or the most prevalent grouping (e.g., ethnicity). Univariate (unadjusted) logistic regression was performed to identify variables associated with intrapartum stillbirth. Variables with *p* <0.2 on univariate analysis were included in adjusted regression models only if they occurred before or at the same timepoint as the independent variable of interest (figure 1), in-order to avoid over adjustment for variables on the same causal pathway.(21) The obstetric response to labouring women who subsequently deliver a stillborn may reflect clinical suspicion of IRA or other obstetric complications instead of being a risk factor for intrapartum stillbirth (figure 1). Hence, we analysed intrapartum obstetric management variables (e.g., delivery mode and place, artificial rupture of membranes and person assisting delivery) separately from other intrapartum variables. Statistical significance for variables associated with stillbirth on the adjusted regression model was defined as *p*<0.05. Missing data were < 1% for all independent variables and available case analysis was used.

**Figure 1.**
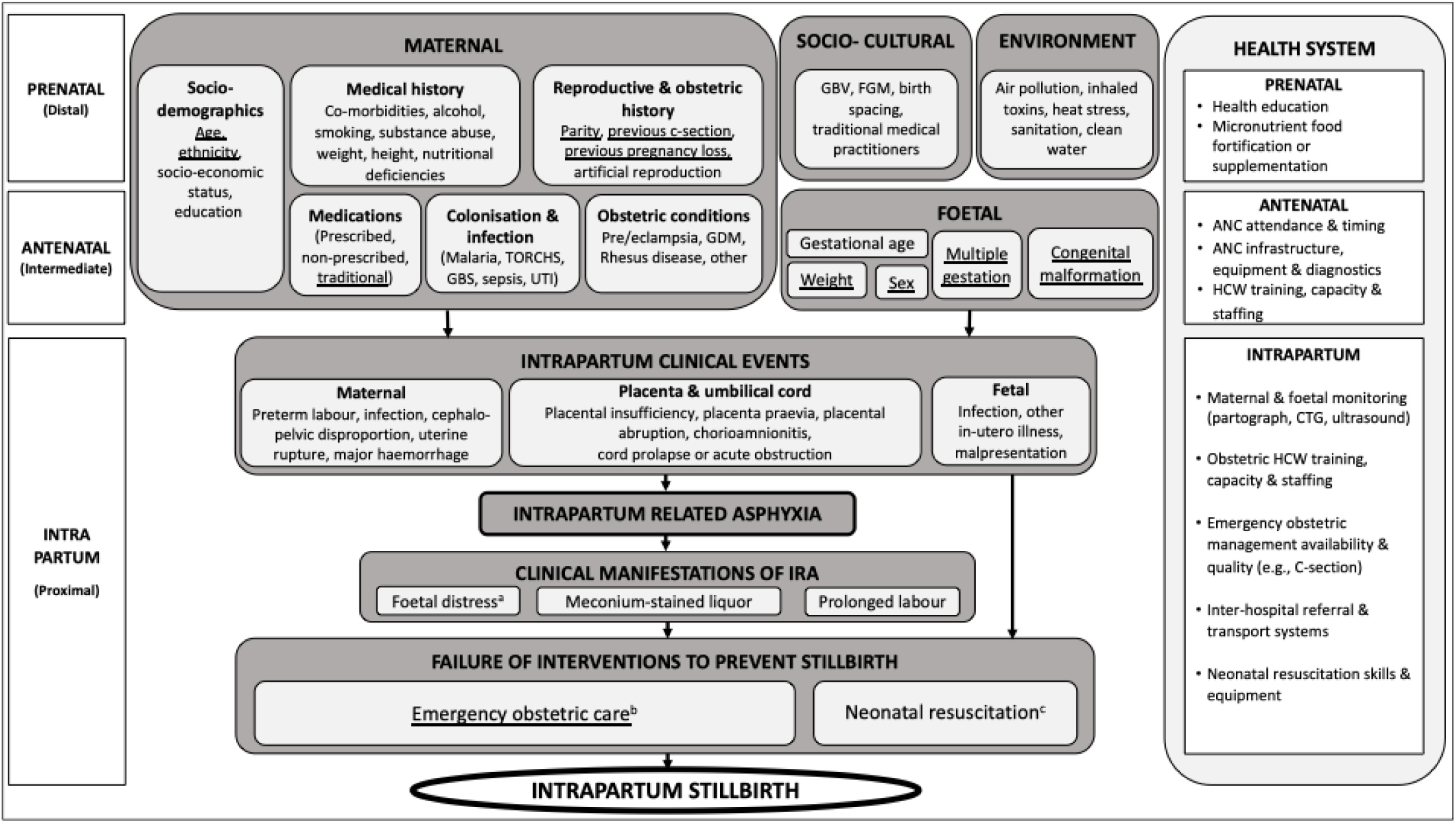
Conceptual framework to understand factors influencing development of intrapartum stillbirth with consideration of health system context (20)(13)) Variables which are underlined were available for the analysis Abbreviations: ANC = Antenatal clinic; CTG = Cardiotocography; FGM: Female genital mutilation; GBS = Group B Streptococcus; GBV = Gender based violence; GDM = Gestational diabetes mellitus; HCW = Health care worker; IRA = Intrapartum related asphyxia; TORCHs = Toxoplasmosis, rubella, CMV, HSV, HIV, syphilis infection; UTI = Urinary tract infection. a. Foetal distress detected as tachycardia, bradycardia or prolonged / unprovoked heart rate decelerations via intermittent auscultation of foetal heartbeat, partograph or cardiotocography. b. Emergency obstetric care interventions include: administration of parenteral antibiotics or anticonvulsants; manual removal of the placenta; assisted delivery by vacuum extraction; caesarean section and blood transfusion. Referral to a higher-level health facility, clinician involvement in delivery process and artificial rupture of membranes were also considered to be interventi ons that may avoid intrapartum stillbirth c. Neonatal resuscitation consists of simulation, bag-mask ventilation, oxygen and chest compressions.

## RESULTS

12,198 total births from 11,983 women were included, of which 55.9% (6,823/12,198) were in The Gambia and 44.1% (5,375/12,198) in Burkina Faso (Figure 2). Baseline characteristics of the study population are shown in Table 1. The intrapartum SBR for the whole cohort was 7.6/1000 total births (93/12,198; 95% CI: 6.2 - 9.3), with 7.3/1000 total births (95% CI: 5.4 - 9.6) in The Gambia and 8.0/1000 total births (95% CI: 5.8 - 10.8) in Burkina Faso (Figure 2).

**Table 1:** Pre-natal, antenatal and intrapartum risk factors for intrapartum stillbirths in a West African cohort of pregnant women.

|  | Total<br>n | Stillbirth<br>n (%) | Live Birth<br>n (%) | Unadjusted regression |  | Adjusted regression |  |
| --- | --- | --- | --- | --- | --- | --- | --- |
|  |  |  |  | OR<br>(95% CI) | p-value | aOR<br>(95% CI) | p-value |
| Pre-natal factors |  |  |  |  |  |  |  |
| Maternal age (years) |  |  |  |  | 0.43 |  |  |
| 20 – 35 | 9546 | 69 (0.7) | 9477 (99.3) | 1 |  |  |  |
| 16 – 19 | 1526 | 12 (0.8) | 1514 (99.2) | 1.09<br>(0.59-2.02) |  |  |  |
| >35 | 1126 | 12 (1.1) | 1114 (98.9) | 1.48<br>(0.80-2.74) |  |  |  |
| Parity |  |  |  |  | 0.01 |  | 0.02 |
| Para 2 | 2245 | 6 (0.3) | 2239 (99.7) | 1 |  | 1 |  |
| Primiparous | 2756 | 27 (1) | 2729 (99) | 3.69<br>(1.52-8.95) |  | 4.16<br>(1.71-10.14)<sup>a</sup> |  |
| Para 3 | 1973 | 16 (0.8) | 1957 (99.2) | 3.05<br>(1.19-7.81) |  | 2.91<br>(1.13-7.47)<sup>a</sup> |  |
| Para ≥4 | 5223 | 44 (0.8) | 5179 (99.2) | 3.17<br>(1.35-7.45) |  | 2.66<br>(1.13-6.29)<sup>a</sup> |  |
| Maternal ethnicity |  |  |  |  | 0.33 |  |  |
| Mossi | 4948 | 39 (0.8) | 4909 (99.2) | 1 |  |  |  |
| Gourounsi | 351 | 3 (0.9) | 348 (99.1) | 1.09<br>(0.34-3.54) |  |  |  |
| Other Burkina Faso | 76 | 1 (1.3) | 75 (98.7) | 1.68<br>(0.23-12.39) |  |  |  |
| Mandinka | 2722 | 15 (0.6) | 2707 (99.4) | 0.70<br>(0.39-1.27) |  |  |  |
| Wolof | 1046 | 5 (0.5) | 1041 (99.5) | 0.60<br>(0.24-1.53) |  |  |  |
| Fula | 1227 | 9 (0.7) | 1218 (99.3) | 0.93<br>(0.45-1.93) |  |  |  |
| Jola | 887 | 11 (1.2) | 876 (98.8) | 1.58<br>(0.81-3.10) |  |  |  |
| Other Gambia | 941 | 10 (1.1) | 931 (98.9) | 1.35<br>(0.67-2.71) |  |  |  |
| Country |  |  |  |  | 0.68 |  |  |
| The Gambia | 6823 | 50 (0.7) | 6773 (99.3) | 1 |  |  |  |
| Burkina Faso | 5375 | 43 (0.8) | 5332 (99.2) | 1.09<br>(0.72-1.64) |  |  |  |
| Previous miscarriage |  |  |  |  | 1.00 |  |  |
| No | 11062 | 85 (0.8) | 10977 (99.2) | 1 |  |  |  |
| Yes | 1135 | 8 (0.7) | 1127 (99.3) | 0.92<br>(0.44-1.90) |  |  |  |
| Previous stillbirth |  |  |  |  | <0.001 |  | <0.001 |
| No | 11897 | 77 (0.7) | 11820 (99.3) | 1 |  | 1 |  |
| Yes | 300 | 16 (5.3) | 284 (94.7) | 8.65 |  | 8.75 |  |
|  |  |  |  | (4.98-15.01) |  | (4.88-15.67)<sup>a</sup> |  |
| **Previous Caesarean section** |  |  |  |  | 0.008 |  | 0.006 |
| No | 12093 | 89 (0.7) | 12004 (99.3) | 1 |  | 1 |  |
| Yes | 104 | 4 (3.9) | 100 (96.1) | 5.40<br>(1.95-14.99) |  | 4.52<br>(1.53-13.33)<sup>a</sup> |  |
| **Antenatal factors** |  |  |  |  |  |  |  |
| **Maternal** |  |  |  |  |  |  |  |
| **Traditional medicine use<sup>b</sup>** |  |  |  |  | 0.008 |  | 0.02 |
| No | 11004 | 91 (0.8) | 10913 (99.2) | 1 |  | 1 |  |
| Yes | 1194 | 2 (0.2) | 1192 (99.8) | 0.20<br>(0.05-0.81) |  | 0.20<br>(0.05-0.81)<sup>c</sup> |  |
| **Pre-labour antibiotics** |  |  |  |  | 0.14 |  | 0.04 |
| No | 12110 | 91 (0.8) | 12019 (99.2) | 1 |  | 1 |  |
| Yes | 87 | 2 (2.3) | 85 (97.7) | 3.11<br>(0.75-12.83) |  | 4.67<br>(1.10-19.86)<sup>c</sup> |  |
| **Pre-labour fever** |  |  |  |  | 0.2 |  |  |
| No | 12167 | 92 (0.8) | 12075 (99.2) | 1 |  |  |  |
| Yes | 29 | 1 (3.5) | 28 (96.5) | 4.69<br>(0.63-34.84) |  |  |  |
| **Pre-labour leaking of liquor** |  |  |  |  | 0.02 |  | 0.10 |
| No | 11769 | 85 (0.7) | 11684 (99.3) | 1 |  | 1 |  |
| Yes | 427 | 8 (1.9) | 419 (98.1) | 2.62<br>(1.26-5.44) |  | 1.94<br>(0.89-4.24)<sup>c</sup> |  |
| **Pre-labour antipyretics** |  |  |  |  | 0.41 |  |  |
| No | 12126 | 92 (0.8) | 12034 (99.2) | 1 |  |  |  |
| Yes | 69 | 1 (1.5) | 68 (98.5) | 1.92<br>(0.26-13.98) |  |  |  |
| **Neonatal** |  |  |  |  |  |  |  |
| **Sex** | 12198 |  |  |  | 0.68 |  |  |
| Female | 5907 | 43 (0.7) | 5864 (99.3) | 1 |  |  |  |
| Male | 6291 | 50 (0.8) | 6241 (99.2) | 1.09<br>(0.72-1.64) |  |  |  |
| **Birth weight (kg)** |  |  |  |  | <0.001 |  | 0.002 |
| Normal (2.5-4.0) | 10870 | 62 (0.6) | 10808 (99.4) | 1 |  | 1 |  |
| Low (<2.5) | 1162 | 22 (1.9) | 1140 (98.1) | 3.36<br>(2.06-5.49) |  | 1.91<br>(1.07-3.40)<sup>c</sup> |  |
| Macrosomia (>4) | 156 | 4 (2.6) | 152 (97.4) | 4.59<br>(1.65-12.78) |  | 4.97<br>(1.74-14.17)<sup>c</sup> |  |
| **Multiple birth** |  |  |  |  | <0.001 |  | <0.001 |
| No (Singleton) | 11764 | 78 (0.7) | 11686 (99.3) | 1 |  | 1 |  |
| Yes (Twin/triplet)<sup>d</sup> | 434 | 15 (3.5) | 419 (96.5) | 5.36<br>(3.06-9.39) |  | 3.53<br>(1.73-7.21)<sup>c</sup> |  |
| **Congenital malformation<sup>e</sup>** |  |  |  |  | 0.68 |  |  |
| No | 11984 | 91 (0.8) | 11893 (99.2) | 1 |  |  |  |
| Yes | 214 | 2 (0.9) | 212 (99.1) | 1.23 |  |  |  |
|  |  |  |  | (0.30-5.03) |  |  |  |
| **Intrapartum factors** |  |  |  |  |  |  |  |
| **Season of birth** |  |  |  |  | 0.27 |  |  |
| Dry (Nov-May) | 7939 | 66 (0.8) | 7873 (99.2) | 1 |  |  |  |
| Wet (Jun-Oct) | 4259 | 27 (0.6) | 4232 (99.4) | 0.76<br>(0.48-1.19) |  |  |  |

Abbreviations: aOR = Adjusted odds ratio; CI = Confidence interval; OR = Odds ratio
a) Adjusted odds ratios adjusted for other pre-natal variables with p-value <0.2 in the unadjusted analysis (parity, previous stillbirth, previous caesarean section); b) Antenatal use of traditional medicine is binary variable, with no information available about type, frequency or timing of use; c) Adjusted odds ratios adjusted for other pre-natal and antenatal variables with p-value <0.2 in the unadjusted analysis (parity, previous still birth, previous caesarean; section, traditional medicine, leaking liquor, antibiotic use, multiple birth, birthweight); d) 428 twins and 6 triplets; e) Easily recognisable congenital malformations

**Figure 2.**
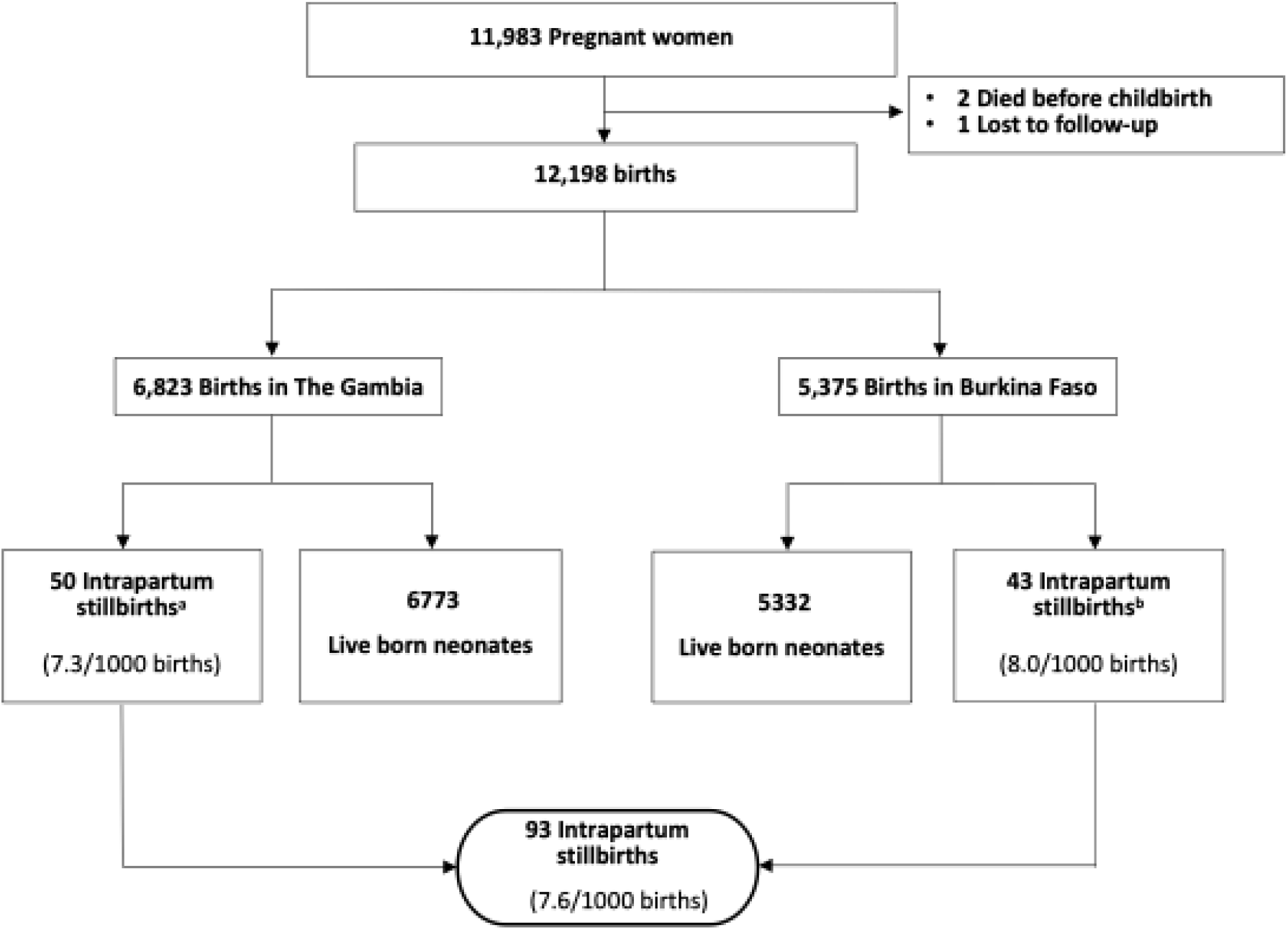
Overview of participants and intrapartum stillbirths in a West African cohort of pregnant women a) 33/50 (66%) fresh stillbirths and 17/50 (33%) macerated stillbirths in The Gambia b) 36/43 (84%) fresh stillbirths and 7/43 (16%) macerated stillbirths in Burkina Faso

### Risk factors for intrapartum stillbirths

Pre-natal risk factors for intrapartum stillbirth in our cohort included: maternal parity, with a 4-fold increase in odds for primiparous women compared with parity-2 women (aOR: 4.16, 95% CI: 1.71 –10.14); women with a history of previous stillbirth (aOR 8.75, 95% CI: 4.88 - 15.67); and women with a history of previous caesarean section (aOR 4.52, 95% CI: 1.53 - 13.33). The only maternal antenatal factor identified on adjusted analysis was antibiotic use before labour (aOR 4.67, 95% CI: 1.10 - 19.86). Foetal factors associated with increased odds of being stillborn included multiple births (mostly twins), (aOR: 3.53, 95% CI: 1.73 - 7.21), and extremes of birth weight (macrosomia, aOR: 4.97, 95% CI: 1.74 –14.17; low birth weight (LBW), aOR: 1.91, 95% CI: 1.07 – 3.40)(Table 1). Use of traditional medicine during pregnancy was a protective factor for intrapartum stillbirth (aOR 0.2, 95% CI: 0.05 - 0.81) (Table 1), although details about type of medicine and timing of administration were not available.

### Association between intrapartum management strategies and intrapartum stillbirth

Delivery by caesarean section (OR 6.55, 95% CI: 3.45-12.45) and a clinician assisting the delivery (OR 6.66, 95% CI: 3.69 – 12.02) had over 6-fold increased odds of stillbirth. Spontaneous rupture of membranes was also associated with intrapartum stillbirth (OR 1.65, 95% CI: 1.05 - 2.59), when compared with women undergoing artificial membrane rupture (Table 2).

**Table 2:**
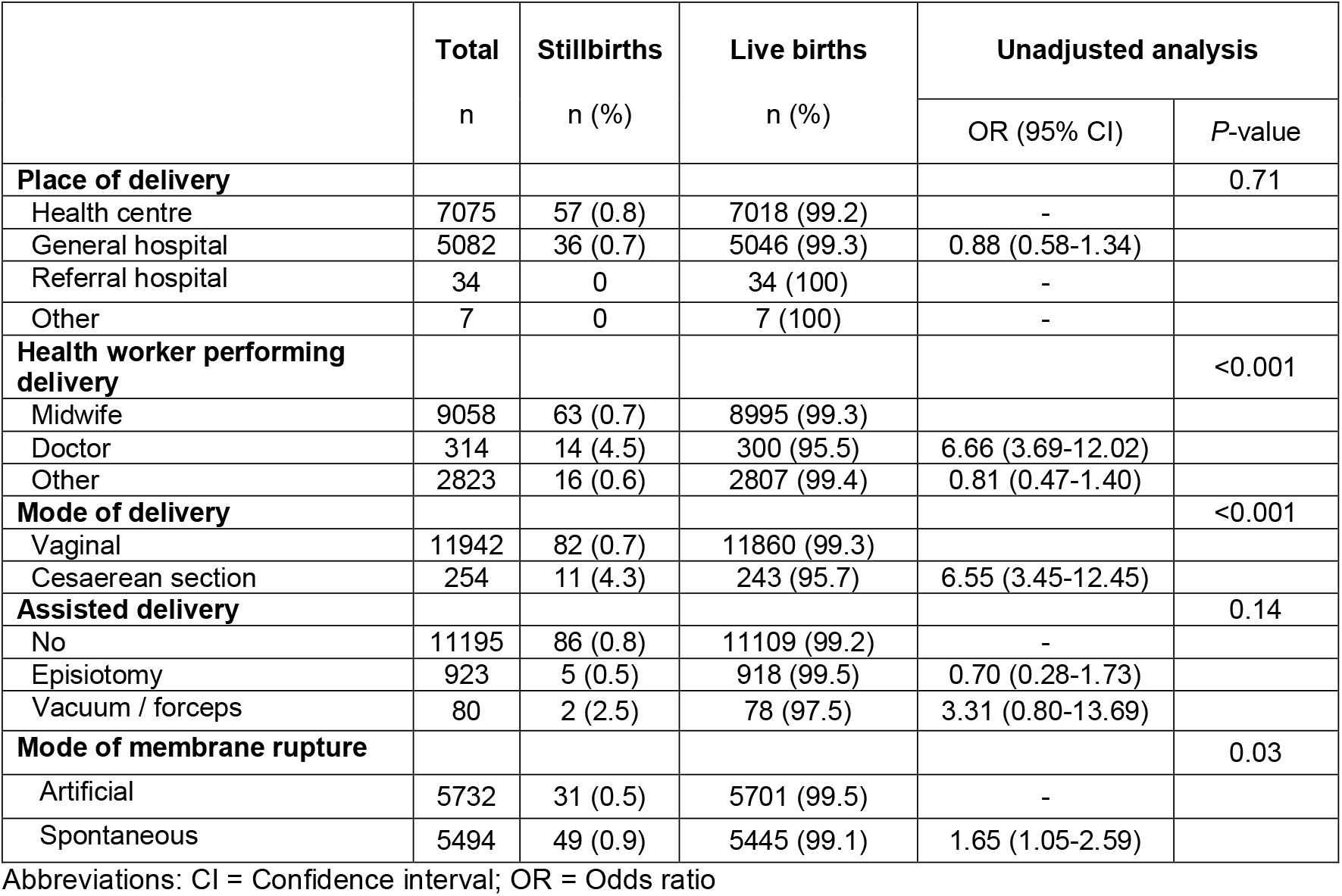
Intrapartum obstetric management factors and intrapartum stillbirths in a West African cohort of pregnant women.

|  | **Total**<br>n | **Stillbirths**<br>n (%) | **Live births**<br>n (%) | **Unadjusted analysis** |  |
| --- | --- | --- | --- | --- | --- |
|  |  |  |  | OR (95% CI) | P-value |
| **Place of delivery** |  |  |  |  | 0.71 |
| Health centre | 7075 | 57 (0.8) | 7018 (99.2) | - |  |
| General hospital | 5082 | 36 (0.7) | 5046 (99.3) | 0.88 (0.58-1.34) |  |
| Referral hospital | 34 | 0 | 34 (100) | - |  |
| Other | 7 | 0 | 7 (100) | - |  |
| **Health worker performing delivery** |  |  |  |  | <0.001 |
| Midwife | 9058 | 63 (0.7) | 8995 (99.3) |  |  |
| Doctor | 314 | 14 (4.5) | 300 (95.5) | 6.66 (3.69-12.02) |  |
| Other | 2823 | 16 (0.6) | 2807 (99.4) | 0.81 (0.47-1.40) |  |
| **Mode of delivery** |  |  |  |  | <0.001 |
| Vaginal | 11942 | 82 (0.7) | 11860 (99.3) |  |  |
| Cesarean section | 254 | 11 (4.3) | 243 (95.7) | 6.55 (3.45-12.45) |  |
| **Assisted delivery** |  |  |  |  | 0.14 |
| No | 11195 | 86 (0.8) | 11109 (99.2) | - |  |
| Episiotomy | 923 | 5 (0.5) | 918 (99.5) | 0.70 (0.28-1.73) |  |
| Vacuum / forceps | 80 | 2 (2.5) | 78 (97.5) | 3.31 (0.80-13.69) |  |
| **Mode of membrane rupture** |  |  |  |  | 0.03 |
| Artificial | 5732 | 31 (0.5) | 5701 (99.5) | - |  |
| Spontaneous | 5494 | 49 (0.9) | 5445 (99.1) | 1.65 (1.05-2.59) |  |

Abbreviations: CI = Confidence interval; OR = Odds ratio

## DISCUSSION

This large multi-centre health-facility based cohort study nested in a clinical trial and conducted across several West African health facilities highlights the substantial risk of intrapartum stillbirth in both rural and urban settings among pregnant women considered to be at low risk for intrapartum complications. We identified several pre-natal and antenatal risk factors for intrapartum stillbirth that can be recognised early in pregnancy, enabling improved triaging with potential for safer deliveries and improved foetal outcomes.

The intrapartum SBR in our study was 7.6 per 1000 total births and was consistent between rural Burkina Faso and urban Gambia. This rate likely underestimates the population-level intrapartum SBR because women with known pregnancy or foetal complications were excluded from our cohort. Our reported rate is half that of a previous pilot study we conducted at one of the two sites (BMCHH) in The Gambia from 2013 to 2014 (15.4 intrapartum stillbirths/1000 total births) which used similar eligibility criteria and definitions,(22) possibly indicating temporal improvements in intrapartum stillbirth rates during this time. Conversely, our rate exceeds the intrapartum SBR reported from African sites in the large AMANHI study (5/1000 total births of > 50,000 deliveries), which included one West African site in Ghana.(23) The AMANHI study highlighted substantial variation in stillbirth rates by country, with Ghana showing one of the highest rates (27/1000 total births), although the timing of stillbirths was indeterminate in 78% of cases, hence the intrapartum SBR may have been underestimated at the Ghanaian AMANI site.(23) Accurate estimation of intrapartum stillbirth rates is challenging due to difficulties in precisely determining and documenting the onset of labour, which is essential for distinguishing between antenatal and intrapartum stillbirths. Our study confirmed the foetal heartbeat prospectively after onset of labour, hence our data only captured intrapartum stillbirths with minimal risk of misclassification.

We identified several risk factors for intrapartum stillbirth, including features of maternal obstetric history (parity, previous stillbirth and Caesarean section), foetal characteristics (multiple gestation, macrosomia, LBW) and maternal antibiotic use prior to labour. These factors have all been identified for intrapartum stillbirths by previous African specific studies,(13, 23, 24) except maternal antibiotic use.

Pre-labour leakage of liquor in our cohort was also associated with stillbirth on unadjusted analysis with a lack of statistical significance on adjusted analysis possibly due to lack of power for this variable. However, other markers of maternal bacterial infection (maternal fever and antipyretic use) were not associated with intrapartum stillbirth in our cohort, so the significance of this finding is not clear. Women with a history of previous stillbirth had 8-fold higher rates of intrapartum stillbirth in our cohort, compared with women who had no experience of late foetal loss. We did not document timing of previous stillbirth, so we cannot comment on whether the previous stillbirth was antenatal or intrapartum. This result is consistent with previous health-facility studies in Nigeria and Ghana(13) and highlights the importance of recognising and preventing recurrent stillbirths in our setting. Asking pregnant women about their obstetric history is potentially feasible, if done in a culturally appropriate and sensitive manner, and could be the basis of a screening tool along with other risk factors to enable early detection and risk stratification of women at high-risk for intrapartum stillbirth. This finding highlights the possibility that stillbirths may be clustered within the same women, with implications for maternal mental health and the need for targeted psychological support.(25, 26) Multiple births (mostly twins) in our cohort had almost four times increased odds of intrapartum stillbirth compared to singletons. The association between twins and intrapartum stillbirth has been extensively reported by African population-based studies,(13, 23, 27, 28) including in West Africa.(13) This underlines the important role of twins in the pathway to intrapartum stillbirth, especially in regions where twinning rates are high, such as West Africa.(29, 30) Women with twin pregnancies who receive antenatal care experience less intrapartum stillbirths compared to those without antenatal care.(29) Hence, our finding supports the need for a programmatic approach to detect twin pregnancies early, in-order to promote antenatal attendance,(28) with close monitoring for maternal complications and optimal foetal growth, and including timely referral to obstetric services for safe delivery.(31) We also identified that extremes of birth weight are associated with 2- to 4-fold increased odds of having an intrapartum stillbirth, with highest increased odds for macrosomia on the adjusted analyses. This is an important addition to the literature, as although foetal macrosomia is associated with increased risk of stillbirth in well-resourced, high-income settings,(32) a recent comprehensive systematic review and meta-analysis of African data identified only weak evidence for macrosomia as a stillbirth risk factor.(13) LBW is a well-recognised risk factor for stillbirths in Africa,(13, 14) including The Gambia where a retrospective cross-sectional study showed 6-fold increase in intrapartum stillbirths for LBW offspring.(33) Our finding of increased intrapartum stillbirth in growth restricted foetuses is limited by the lack of gestational age data for our cohort, as prematurity is strongly associated with stillbirths with the highest risk for premature and small-for-gestational age phenotypes.(34) We did not identify any association between either maternal age or congenital malformations and intrapartum stillbirths, which have been reported previously by West African studies,(13) although this may be related to limited power for these variables in our cohort.(13)

Our finding of increased intrapartum stillbirths following emergency caesarean section delivery and clinician presence at birth is not likely to be causative but may indicate that complicated deliveries and signs of foetal distress were recognised and attempts made to expedite delivery, albeit not timely enough to prevent foetal loss. Our finding that women were more likely to experience stillbirth if their cervical membrane had spontaneously ruptured is consistent with an Ethiopian health-facility based case-control study which identified membrane rupture at time of admission as a determinant of intrapartum stillbirth.(24) This may indicate prolonged rupture of membranes, known to increase stillbirth risk via infectious aetiologies, (13, 35) or may represent a protective effect from artificial rupture of membranes, an established obstetric strategy used to hasten delivery. However, we recommend caution in over interpreting this finding as the data was based on retrospective recall by mothers following delivery and may be subject to recall bias.

The findings of this study are generalisable to pregnant women considered to be at low risk for intrapartum complications in resource-limited West African health facilities but may not be applicable to non-research populations at higher-risk or within better-resourced health systems. Furthermore, the study was constrained by a limited choice of independent variables due to the post-hoc design using pre-existing data. There are important gaps in the data, including variables related to maternal morbidities (e.g., hypertension, confirmed infection, antepartum and postpartum haemorrhage) and obstetric complications (e.g., polyhydramnios, chorioamnionitis, uterine abruption, prolonged labour and malpresentation), which have previously been associated with intrapartum stillbirths.(13, 23) Additionally, process variables regarding health system factors such as health worker capacity, referral pathways and quality of intrapartum care including use of partographs or intermittent auscultation for timely detection of foetal distress was lacking(35) and should be included in future research. Detailed data on traditional medicine usage (e.g., type, frequency, duration of use) would have enabled interpretation of our finding that antenatal traditional medicine use appears protective against intrapartum stillbirth. This observation contrasts with other West African data suggesting that herbal use during pregnancy increases the risk of intrapartum stillbirth, (36) and further research is warranted.

## CONCLUSION

This study underlines the persistently high rates of intrapartum stillbirths in two West African countries among pregnant women who may previously have been considered low risk for intrapartum complications or stillbirth. Early identification of risk factors in pregnant women (first pregnancy, previous stillbirth or caesarean section, twins or foetal macrosomia/LBW), combined with early and regular antenatal care with risk stratification and planning for safe health facility delivery, holds significant promise for preventing intrapartum stillbirths in West Africa.

## Data Availability

Data are not publicly available. Qualified researchers may request access with the Gambia Government/MRC Joint Ethics Committee. The review process and release of data will be facilitated by MRC Unit The Gambia (http://www.mrc.gm/) through the Head of Governance. Access will not be unduly restricted.

## ABBREVIATIONS

aOR: Adjusted odds ratio
BMCHH: Bundung Maternal and Child Health Hospital
CI: Confidence interval
CMA: Centre médical avec antenne chirurgicale
HIC: High-income countries
HIV: Human Immunodeficiency Virus
IRA: Intrapartum related asphyxia
LBW: Low birth weight
LMIC: Low- and middle-income countries
OR: Odds ratio
SBR: Stillbirth rate
SHC: Serekunda Health Centre
SSA: sub-Saharan Africa

## DECLARATIONS

### Ethics approval and consent to participate

The trial was approved by The Gambia Government/MRCG (Medical Research Council Unit The Gambia) Joint Ethics Committee, the Comité d’Ethique pour la Recherche en Santé (CERS) and the Ministry of Health of Burkina Faso, and the LSHTM Ethics Committee. All women provided written informed consent and were free to withdraw from the study at any stage.

### Consent for publication

Not applicable

### Declaration of competing interests

The authors declare that they have no competing interests.

### Funding

The PregnAnZI-2 trial was funded by a grant from the UKRI under the Joint Global Health Trial Scheme (JGHT) (ref: MC_EX_MR/P006949/1) and the Gates Foundation (Ref:OPP1196513). The publication of this supplement was funded in whole by the Gates Foundation. The funders and study sponsor (MRCG) had no role in the study design, collection, analysis or interpretation of data, writing of article nor the decision to submit for publication.

### Authors’ contributions

AR conceptualised this study with input from UDA, HB, HT, CB, JDB and BC. Data collection was co-ordinated in The Gambia by BC with UNN, NB, SG, CJ, MD, and FS contributing to recruitment and data collection. The Burkina Faso data collection team included GJWN, EYS, and AMS with co-ordination by JDB. YN and EN curated and cleaned the data with input from BC, JDB, UNN, TR, SG, CJ, MD, FS, GJWN, EYS, and AMS. The conceptual framework was developed by FS, NB and HB. CB performed the analysis with full access to the data. FS drafted the initial manuscript with input from HB, TR, and AR. All authors read and approved the final version. AR gave oversight to the work as guarantor and accepts full responsibility for the finished work and controlled the decision to publish.

## Acknowledgements

Firstly, we acknowledge all the women, their newborns and families who participated in the PregnANZI-2 trial and made this study possible. We are thankful to the health facility health workers and other staff at the PregnAnZI-2 trial sites for their collaboration and contributions. We appreciate the collaboration with the Ministries of Health in The Gambia and Burkina Faso to enable the study to be conducted. We thank all research and research support personnel at both MRC Unit The Gambia at LSHTM and the Clinical Research Unit of Nanoro (CRUN) in Burkina Faso, without which the study would not have been possible.

## Notes

### Competing Interest Statement

The authors have declared no competing interest.

### Clinical Trial

NCT03199547

### Author Declarations

Ethical approval for this work was provided by The Gambia Government/MRCG (Medical Research Council Unit The Gambia) Joint Ethics Committee, the Comite de Ethique pour la Recherche en Sante (CERS) and the Ministry of Health of Burkina Faso, and the LSHTM Ethics Committee.

